# Acceptability and implementation considerations of the patient-collected rectal swab for sexually transmitted infections testing among men who have sex with men and transgender women in Kigali, Rwanda

**DOI:** 10.1101/2024.09.30.24314599

**Authors:** Jean Olivier Twahirwa Rwema, Nneoma E. Okonkwo, Matthew M. Hamill, Carrie E. Lyons, Neia Prata Menezes, Jean Damascene Makuza, Julien Nyombayire, Gallican Nshogoza Rwibasira, Aflodis Kagaba, Patrick Sullivan, Susan Allen, Etienne Karita, Stefan D. Baral

**Affiliations:** Department of Epidemiology, Key Populations Program, Center for Public Health and Human Rights, Johns Hopkins Bloomberg School of Public Health, Baltimore, MD; School of Medicine, Johns Hopkins University, Baltimore, MD; Division of Infectious Diseases, Johns Hopkins School of Medicine, Baltimore, MD; Projet San Francisco, Kigali, Rwanda; Rwanda Biomedical Center, HIV and AIDS division, Kigali, Rwanda; School of Population and Public Health, University of British Columbia, Vancouver, BC, Canada; Health Development Initiative, Kigali, Rwanda; Department of Epidemiology, Rollins School of Public Health, Emory University, Atlanta, GA, USA

**Keywords:** sexually transmitted infections, patient-collected samples, rectal STI testing, men who have sex with men, transgender women, Kigali, Rwanda

## Abstract

**Background:** Rectal sexually transmitted infections (STI) are prevalent among men who have sex with men (MSM) and transgender women (TGW). Self-collection of rectal specimens is widely used globally, but limited data exist on its implementation in Africa. We report experiences of MSM/TGW self-collecting rectal STI specimens in Kigali.

**Methods:** From March to August 2018, 738 MSM/TGW were recruited in a cross-sectional study using respondent-driven sampling in Kigali. We tested for *Neisseria gonorrhea* and *Chlamydia trachomatis* using the Cepheid GeneXpert CT/NG platform on self-collected rectal swabs. Likert scales assessed self-collection difficulty and comfort. Ordered logistic regression analyses were performed to characterize factors associated with self-collection difficulty.

**Results:** Overall, 14%(106) identified as TGW. In total, 78%(577) found rectal swab self-collection easy/very easy, while 15%(108) found it difficult/very difficult. Most, 92%(679), were comfortable/very comfortable with the test, and 98%(730) said they would repeat the test in the future. In multivariable RDS-adjusted analyses, discomfort with the swab was positively associated with difficulty in self-collection (adjusted cumulative odds ratios ((aCOR): 7.9(95%CI: 4.9-12.8)) and having a prevalent rectal STI (aCOR: 4.19, 95%CI: 2.02-8.72) was significantly associated with difficulty.

Furthermore, 10%(75) of rectal swabs returned indeterminate results (65 invalid results and 10 errors).

**Conclusion:** Most MSM/TGW found self-collection of rectal swabs easy, comfortable, and collected valid samples. Self-collection of rectal swabs could be used alongside clinic- and community-based STI testing to improve diagnosis and treatment in Rwanda. However, measures to optimize sample collection and processing are needed to reduce the cost and clinical implications of indeterminate results

## Introduction

The incidence and prevalence of sexually transmitted infections (STI) continue to increase globally ^1^. STI are a major public health issue, as they are associated with increased risk of HIV acquisition and transmission, adverse sexual and reproductive health outcomes, and increased years lived with disability ^1-3^. Men who have sex with men (MSM) and transgender women (TGW) are at higher risk of STI compared to aged-matched non-MSM/TGW because of intersecting individual, network, and structural factors ^4-6^. STI prevalence among MSM/TGW in our recent study in Kigali was high, with approximately one in five MSM/TGW having an STI infection ^7^. Furthermore, 74% of participants diagnosed with an STI in this study did not exhibit any symptoms, and nearly a third of *Chlamydia trachomatis* (CT) infections and over half of *Neisseria gonorrhoeae* (NG) infections were rectal ^7^. Other studies have demonstrated the high burden of rectal STI in MSM/TGW, and that urogenital-only-testing misses the vast majority of extragenital infections ^8^.

Current evidence from high-income settings shows that self-collection of biological samples for STI testing, including rectal swabs, is highly acceptable to users, and performs similarly to provider-collected specimens ^9-12^. Additionally, offering self-collection of samples as an option for STI testing can result in increased testing uptake and detection of STI in clinical settings ^13,14^. Finally, self-collection of specimens can be used to support community and home-based testing to reach MSM/TGW with limited access to healthcare settings ^15,16^. However, data about self-collection from countries across SSA and other low- and middle-income countries (LMICs) are limited.

As in many countries across sub-Saharan Africa (SSA), STI programming in Rwanda, including optimal prevention and treatment services, is hampered by several factors ^17,18^. Mainly, STI management and surveillance in SSA mostly rely on syndromic case management, which results in under diagnosis, misdiagnosis, and suboptimal treatment because of its low sensitivity and specificity, which may increase sequelae and antibiotic resistance ^19-21^. MSM/TGW face additional challenges due to socio-structural factors including stigma from communities and healthcare professionals which result in low utilization of healthcare services ^17,18^. Health care provision is further limited because healthcare space rarely considers the particular needs of sexual and gender minorities. For example, extra genital infections (rectal and pharyngeal) are seldom screened for in clinical settings across SSA. Finally, refusal of rectal STI screening among MSM/TGW in SSA has been reported even when individuals present at clinics ^22^. A recent study among MSM/TGW in South Africa reported high refusal of clinician-collected rectal STI screening despite universal acceptance of self-collected genital testing ^22^. Refusal of clinical examination by sexual and gender minorities has been documented previously and has been linked in part to a lack of trust in healthcare providers’ knowledge of sexual and gender minorities’ needs ^23^. Stigma associated with sexual behaviors and fear of disclosing same-sex relationships are also barriers to healthcare utilization among MSM/TGW ^24,25^. As a result, several strategies have been designed to support STI programming in clinical and community settings such as self-collection of biological samples, including rectal swabs ^26^.

In this study, we described experiences of MSM/TGW using self-collected rectal swabs for CT/NG testing in the context of a behavioral and biological survey conducted in Kigali, Rwanda. We further evaluated factors associated with self-reported difficulty in performing rectal swab self-collection and discuss potential implementation challenges of integrating self-collection of biological specimens especially rectal swabs to support clinical-, community-, and home-based STI testing in Rwanda.

## Materials and Methods

### Study setting and procedures

A cross sectional behavioral and biological assessment among MSM and TGW in Kigali, was conducted from March-August 2018 using respondent driven sampling (RDS) – a method that leverages peer networks to recruit members of marginalized populations ^32^. The study procedures and methods have been previously published ^7,17^. In summary, recruitment was initiated with two recruitment seeds, a third seed was added to enhance recruitment in older MSM/TGW networks. Seeds are individuals who begin the recruitment chains in RDS studies. Eligible participants were individuals assigned the male sex at birth, aged ≥18 years, living in Kigali, and who reported anal sex with another man in the 12 months preceding study enrolment. After eligibility screening, and providing signed informed consent, participants underwent biological testing for HIV, Syphilis, CT, and NG and completed an interviewer-administered questionnaire. Before the interview, a blood sample was collected for each participant for HIV and Syphilis testing. Furthermore, all participants self-collected urine and rectal swabs for CT and NG testing. Further details on laboratory procedures for HIV and STI testing in this study as well as prevalence proportions of these infections have been previously published ^7,17^.

### Rectal swabs collection for CT/NG testing

All eligible participants received verbal instructions on self-collection of urine and rectal swabs. These instructions were provided by the nurses administering the survey who had been trained on self-collection techniques and healthcare needs of MSM/TGW. After receiving the instructions, participants collected their specimens in a bathroom facility at the study site that was solely used for specimen collection purposes. For rectal swabbing, participants were instructed to insert the swab into the rectum and rotate the swab for 5-10 seconds. The swab was then placed in a capped tube. The samples were returned to their interviewer and transported to the onsite laboratory before initiating the interview. All samples were refrigerated and transported to Projet San Francisco (PSF) laboratory in Kigali for processing daily. All rectal specimens were tested for CT and NG using the CT/NG Cepheid GeneXpert platform (Solna, Sweden) within 24 – 36 hours of collection. Participants who tested positive for any of the STI screened for in the study were treated free of charge at the study site according to the Rwanda national guidelines ^17^. When a rectal swab result was indeterminate (defined as invalid or error), the sample was retested. If the test remained indeterminate after retesting, participants were invited to the study site to provide a second rectal swab. The study was approved by the Institutional review board of Emory University and the Rwanda National Ethics committee: IRB00089599.

## Measures

### Acceptability of rectal swab self-collection among participants

The survey included questions assessing participants’ experiences about the self-collection of rectal swabs since this is not routinely offered in Rwanda. Specifically, questions were asked to assess acceptability of rectal swab self-collection among participants.

To assess difficulty, participants were asked the following question: “ *How easy or difficult was it for you to collect your STI testing samples with that swab?*” and for comfort: *“How comfortable or uncomfortable was it using the rectal swab or the swab that you used on the inside of your butt?”* Five item Likert scales were used to assess the level of difficulty (from very difficult to very easy) and comfort (very uncomfortable to very comfortable) of the rectal swabbing process.

The outcome variable was difficulty encountered during the self-collection of the rectal swab. Participants were also asked whether they would be willing to retake a rectal swab for STI testing in the future.

### Other measures of interest

Several factors were hypothesized to be associated with experiencing difficulty with rectal swab self-collection. These included sociodemographic, behavioral, and health-related factors. The sociodemographic characteristics considered were age, education level, income level, occupation, marital status, sexual orientation, and gender identity. Gender identity was assessed using the two-step gender assessment questionnaire and individuals were categorized as cisgender MSM or TGW ^27^. Sexual orientation was classified as homosexual or gay, bisexual, and heterosexual based on survey responses. Behavioral factors included the number of sexual partners and the type of sexual activity respondents engaged in. Health-related, and biological factors included markers of poor mental health, self-reported STI history within the previous 12 months and HIV/STI status. Mental health status was assessed using the Patient Health Questionnaire 9 and a cutoff of > 10 was used for depression ^28^. HIV and STI status were based on testing performed in the study ^17,29^. Finally, the proportion of indeterminate test results, defined as invalid or error, and repeat testing outcome data were collected.

### Statistical analyses

For analysis, the five-item Likert scales for difficulty and comfort were transformed into three-item scales based on the distribution of participants’ responses. The three category variables were (Easy/Very Easy – Neutral – Difficult/ Very Difficult) for difficulty and (Comfortable/Very Comfortable – Neutral - Uncomfortable/Very Uncomfortable) for comfort. We calculated proportions for demographic, behavioral, mental health, and biologic variables by difficulty implementing the rectal swab (Table 1).

**Table 1.** Men who have sex with men and transgender women’s experiences with self-collected rectal swabs in Kigali, Rwanda 2018 (N=738)

|  | Difficulty in self-collecting rectal swab |  |  | χ<sup>2</sup> P value |
| --- | --- | --- | --- | --- |
|  | Easy<br>% (N) | Neutral<br>% (N) | Difficult<br>% (N) |  |
| **Characteristics** |  |  |  |  |
| **Age in years** |  |  |  |  |
| 18-24 | 80.9 (272) | 4.8 (16) | 14.3 (48) | 0.222 |
| 25-34 | 75.7 (225) | 9.4 (27) | 14.9 (44) |  |
| Over 35 | 76.9 (80) | 7.7 (8) | 15.4 (16) |  |
| **Education** |  |  |  |  |
| Primary level or less | 82.9 (175) | 7.6 (15) | 9.5 (20) | 0.132 |
| Some secondary | 77.9 (190) | 6.9 (17) | 15.2 (37) |  |
| Secondary or above | 75.3 (212) | 6.7 (19) | 18 (51) |  |
| **Monthly Income (Frw)** |  |  |  |  |
| Less than 50,000 | 80.1 (386) | 6.2 (31) | 13.7 (66) | 0.09 |
| 50-100,000 | 78.5 (140) | 8.5 (15) | 13 (23) |  |
| Over 100,000 | 67.1 (52) | 7.9 (6) | 25 (19) |  |
| **Self-reported gender identity** |  |  |  |  |
| Cis MSM | 77.0 (486) | 7.1 (45) | 15.9 (100) | **0.064** |
| Transgender | 86.7 (91) | 5.7 (6) | 7.6 (8) |  |
| **Self-reported sexual orientation** |  |  |  |  |
| Gay or Homosexual | 79.2 (376) | 6.3 (30) | 14.5 (69) | 0.404 |
| Bisexual | 77.0 (175) | 7.1 (16) | 15.9 (36) |  |
| Heterosexual | 76.5 (26) | 14.7 (5) | 8.8 (3) |  |
| **Sex with women** |  |  |  |  |
| Never | 83.5 (156) | 4.8 (8) | 11.7 (22) | **0.027** |
| Yes, but not in the last 12 months | 79.2 (221) | 5 (14) | 15.8 (44) |  |
| Yes, in the last 12 months | 73.7 (199) | 10.7 (29) | 15.6 (42) |  |
| **Sexual positioning (Ever)** |  |  |  |  |
| Insertive only | 73.3 (200) | 7.7 (21) | 19 (52) | **0.033** |
| Both | 80.5 (335) | 6.5 (27) | 13 (54) |  |
| Receptive only | 89.8 (44) | 6.1 (3) | 4.1 (2) |  |
| **Mental Health status** |  |  |  |  |
| No depressive symptoms | 79.9 (544) | 6.6 (45) | 13.5 (92) | **0.002** |
| Depressed | 60.0 (33) | 10.9 (6) | 29.1 (16) |  |
| **Biological** |  |  |  |  |
| **Self-reported STI history in the 12 months prior to the study** |  |  |  |  |
| No | 80.3 (473) | 6.5 (38) | 13.2 (78) | **0.04** |
| Yes | 70.8 (104) | 8.8 (13) | 20.4 (30) |  |
| **HIV infection** |  |  |  |  |
| Negative | 78.4 (518) | 6.4 (42) | 15.3 (101) | 0.09 |
| Positive | 78.4 (58) | 12.2 (9) | 9.5 (7) |  |
| **Current STI infection** |  |  |  |  |
| Negative | 79.2 (469) | 6.4 (39) | 14.4 (84) | 0.471 |
| Positive | 74.8 (110) | 8.8 (13) | 16.4 (24) |  |
| **Urethral STI** |  |  |  |  |
| Negative | 78.2 (527) | 6.7 (45) | 15.1 (102) | 0.538 |
| Positive | 81.7 (49) | 8.3 (5) | 10 (6) |  |
| **Rectal STI** |  |  |  |  |
| Negative | 79.7 (546) | 6.1 (42) | 14.2 (97) | **0.004** |
| Positive | 61.2 (30) | 16.3 (8) | 22.5 (11) |  |

We used Chi-squared comparisons to compare the difficulty and comfort of the rectal swab across the different variables of interest (Tables 1 & 2). Furthermore, chi-squared comparisons were also computed to assess differences in indeterminate results by different sociodemographic, behavioral, and biological factors. (Supplement Table 1).

To assess factors associated with experiencing difficulty with the rectal swab self-collection, we first fitted simple ordered logistic regression models (proportional odds model) to assess the association between demographic, behavioral, health, and biological factors with self-reported difficulty. For each model, cumulative odds ratios (COR) and 95% confidence intervals (95%CI) were computed. The final multivariable regression model was constructed using variables that showed an association with the outcome at P <0.1 in the bivariable analyses. A Brant test was used to check the proportional odds assumption of the ordered logistic model. The Brant test P value was 0.5 suggesting that the proportional odds assumption was not violated. All analyses were computed using Stata 15 (StataCorp, College Station, Texas, USA) statistical software. RDS adjustments were performed using RDS II estimator.

## Results

In total, 738 participants were recruited with a mean age of 27.4 years (range: 18-68) and 14%(RDS adjusted proportion: 10.6 (95%CI: 7.8-13.5) identified as TGW. One in five participants 20% (RDS-adjusted 16.7% (95%CI: 13.2-20.2) was diagnosed with an STI infection (CT, NG, or syphilis). Chlamydia prevalence (genital + rectal) was 9.1% (RDS-adjusted: 6.1% (95% CI: 3.9-8.4)) and the prevalence of gonorrhea (genital + rectal) was 8.8% (RDS-adjusted: 7.1% (95%CI: 4.9-9.2). Overall, 52% of NG infections and 27% of CT infections were rectal only.

### MSM/TGW experiences with rectal swab self-collection

In total, 78% (577) reported that rectal swab self-collection was easy/very easy, 7% (51) were neutral and 15% (108) found it difficult/very difficult. Most, 92% (678), reported the procedure to be comfortable or very comfortable, 3% (26) were neutral and 5% (34) found it uncomfortable/very uncomfortable. Nearly all participants, 98.9% (727), reported they would be willing to take a rectal swab for STI testing in the future. Overall, TGW were less likely to report experiencing difficulty self-collecting the swab compared to cis-MSM (8% vs 16%).

Furthermore, participants who reported receptive anal sex were significantly less likely to report difficult with self-collection compared to those who never engaged in receptive anal sex (4% vs 19%)). Regarding biological factors, individuals with a history of STI symptoms in the 12 months preceding the study (20% vs 13%) and those with a rectal infection (22% vs 14%) reported higher difficulty compared to those who did not. The proportions of participants reporting difficulty collecting the rectal swab across different sociodemographic, behavioral, and biological factors are reported in Table 1.

Individuals who reported more discomfort collecting the swab reported more years of education compared to those with less education (χ2 P:0.003). Additionally, individuals with prior history of STI symptoms in the previous 12 months (χ2 P: 0.01) and those with depression (χ2 P: 0.01) reported higher levels of discomfort compared to those without. There were also significant differences in comfort with the rectal swab by self-reported sexual practices. For instance, of the MSM/TGW who reported engaging in receptive anal sex only, none reported being uncomfortable with the rectal swab self-collection while 7% of MSM reporting insertive anal sex only reported discomfort (Table 2).

**Table 2.** Comfort with rectal swab self-collection for STI testing among men who have sex with men and Transgender women in Kigali, Rwanda (N=738)

2. Comfort with rectal swab self-collection for STI testing among men who have sex with men and Transgender women in Kigali, Rwanda (N=738)
|  | Comfort with self-collecting rectal swab |  |  | χ<sup>2</sup> P value |
| --- | --- | --- | --- | --- |
|  | Comfortable<br>% (N) | Neutral<br>% (N) | Uncomfortable<br>% (N) |  |
| **Characteristics** |  |  |  |  |
| **Age in years** |  |  |  |  |
| 18-24 | 91.8 (310) | 14 (4.1) | 4.1 (14) | 0.843 |
| 25-34 | 91.6 (272) | 3 (3.0) | 5.4 (16) |  |
| Over 35 | 93.3 (97) | 2.9 (3) | 3.8 (4) |  |
| **Education** |  |  |  |  |
| Primary level or less | 97.1 (204) | 1 (2) | 1.9 (4) | **0.003** |
| Some secondary | 90.9 (222) | 5.3 (13) | 3.7 (9) |  |
| Secondary or above | 88.7 (252) | 3.8 (11) | 7.4 (21) |  |
| **Monthly Income (Frw)** |  |  |  |  |
| Less than 50,000 | 93 (449) | 3.3 (16) | 18 (3.7) | 0.117 |
| 50-100,000 | 91 (162) | 4.5 (8) | 4.5 (8) |  |
| Over 100,000 | 87 (67) | 2.6 (2) | 10.4 (8) |  |
| **Self-reported gender identity** |  |  |  |  |
| Cis MSM | 91.3 (578) | 4.1 (26) | 4.6 (29) | 0.107 |
| Transgender | 95.2 (100) | 0 (0) | 4.8 (5) |  |
| **Self-reported sexual orientation** |  |  |  |  |
| Gay or Homosexual | 92.2 (439) | 3.4 (16) | 4.4 (21) | 0.231 |
| Bisexual | 91.6 (208) | 4.4 (10) | 3.9 (9) |  |
| Heterosexual | 31 (88.6) | 0 (0) | 4 (11.4) |  |
| **Sex with women** |  |  |  |  |
| Never | 93 (174) | 3.2 (6) | 3.8 (7) | 0.889 |
| Yes, but not in the last 12 months | 91.8 (256) | 3.9 (11) | 4.3 (12) |  |
| Yes, in the last 12 months | 91.1 (247) | 3.4 (9) | 5.6 (15) |  |
| **Mental Health status** |  |  |  |  |
| No depressive symptoms | 92.7 (633) | 3.2 (22) | 4.1 (28) | **0.017** |
| Depressed | 81.9 (45) | 7.3 (4) | 10.9 (6) |  |
| **Sexual positioning (Ever)** |  |  |  |  |
| Receptive only | 88.6 (242) | 4.8 (13) | 6.6 (18) | **0.063** |
| Both | 93 (387) | 3.1 (13) | 3.9 (16) |  |
| Insertive only | 49 (100) | 0 (0) | 0 (0) |  |
| **Biological** |  |  |  |  |
| **Self-reported STI history in the 12 months prior to the study** |  |  |  |  |
| No | 92.1 (544) | 4.2 (25) | 3.7 (22) | **0.01** |
| Yes | 91.1 (134) | 0.7 (1) | 8.2 (12) |  |
| **HIV infection** |  |  |  |  |
| Negative | 91.7 (608) | 3.8 (25) | 4.5 (30) | 0.541 |
| Positive | 93.2 (69) | 1.4 (1) | 5.4 (4) |  |
| **STI infection** |  |  |  |  |
| Negative | 92.2 (545) | 3.4 (20) | 4.4 (26) | 0.787 |
| Positive | 90.5 (133) | 4.1 (6) | 5.4 (8) |  |
| **Urethral STI** |  |  |  |  |
| Negative | 91.9 (621) | 3.4 (23) | 4.7 (32) | 0.884 |
| Positive | 93.3 (56) | 3.3 (2) | 3.4 (2) |  |
| **Rectal STI** |  |  |  |  |
| Negative | 92.2 (634) | 3.4 (23) | 4.4 (30) | 0.449 |
| Positive | 87.7 (43) | 4.1 (2) | 8.2 (4) |  |

### Factors associated with difficulty with the rectal swab

Results of the bivariable analyses are reported in table 3. In the multivariable RDS-adjusted analyses, discomfort with the test was significantly associated with difficulty with self-collection of rectal swabs. The adjusted cumulative odds of experiencing difficulty with the rectal swab were 8 times higher, (aCOR): 5.37 (95%CI: 2.39-12.09), for participants who were uncomfortable with the test compared to those without discomfort. Additionally, participants with a prevalent rectal STI had four times the cumulative odds of experiencing difficulty collecting the swab compared to those who did not aCOR: 4.19(95%CI: 2.02-8.72) (Table 3).

**Table 3.** Factors associated with difficulty in self-collecting rectal swabs for STI testing among men who have sex with men and transgender women in Kigali, Rwanda.

3. Factors associated with difficulty in self-collecting rectal swabs for STI testing among men who have sex with men and transgender women in Kigali, Rwanda
|  | Unadjusted<br>cumulative OR | p-value | Adjusted<br>Cumulative OR<br>(95%CI) | p-value | RDS- adjusted<br>Cumulative OR<br>(95%CI) | p-value |
| --- | --- | --- | --- | --- | --- | --- |
| **Discomfort with rectal swabbing** |  |  |  |  |  |  |
| Self-reported discomfort | **8.61( 5.45-13.60)** | **0.0001** | **7.91(4.91-12.76)** | **0.0001** | **5.37(2.39-12.09)** | **0.0001** |
| **Sociodemographic characteristics** |  |  |  |  |  |  |
| **Age in years** |  |  |  |  |  |  |
| 18-24 | Ref |  |  |  |  |  |
| 25-34 | 1.31(0.90-1.92) | 0.156 |  |  |  |  |
| Over 35 | 1.25(0.74--2.13) | 0.398 |  |  |  |  |
| **Self-reported gender identity** |  |  |  |  |  |  |
| Cis MSM | Ref |  | Ref |  | Ref |  |
| Transgender | **0.51(0.28-0.91)** | **0.024** | 0.54(0.26-1.13) | 0.103 | 0.44(0.16-1.16) | 0.097 |
| **Self-reported sexual orientation** |  |  |  |  |  |  |
| Gay or Homosexual | Ref |  |  |  |  |  |
| Bisexual | 1.12(0.77-1.63) | 0.557 |  |  |  |  |
| Heterosexual | 1.02(0.45-2.28) | 0.959 |  |  |  |  |
| **Education** |  |  |  |  |  |  |
| Primary level or less | Ref |  | Ref |  | Ref |  |
| Some secondary | 1.41(0.88-2.25) | 0.144 | 1.19(0.69-2.06) | 0.524 | 1.47(0.74-2.94) | 0.269 |
| Secondary or above | **1.65(1.05-2.57)** | **0.028** | 1.15(0.65-2.01) | 0.632 | 1.28(0.54-3.05) | 0.574 |
| **Monthly Income (Frw)** |  |  |  |  |  |  |
| Less than 50,000 | Ref |  | Ref |  | Ref |  |
| 50-100,000 | 1.06(0.69-1.61) | 0.792 | 0.97(0.59-1.58) | 0.899 | 0.94(0.49-1.79) | 0.845 |
| Over 100,000 | **1.96(1.16-3.29)** | **0.011** | 1.48(0.78-2.79) | 0.228 | 1.81(0.62-5.31) | 0.277 |
| **Behavioral Characteristics** |  |  |  |  |  |  |
| **Sex with women** |  |  |  |  |  |  |
| Never | Ref |  |  |  | Ref |  |
| Yes, but not in the last 12 months | 1.35(0.83-2.17) | 0.224 | 1.27(0.74-2.21) | 0.383 | 1.33(0.54-3.33) | 0.534 |
| Yes, in the last 12 months | **1.72(1.07-2.75)** | **0.023** | 1.37(0.79-2.36) | 0.251 | 1.35(0.60-3.01) | 0.468 |
| **Sexual positioning** |  |  |  |  |  |  |
| Receptive only | **Ref** |  |  |  | Ref |  |
| Both | 2.21(0.85-5.73) | 0.103 | 1.35(0.47-3.88) | 0.567 | 1.13(0.36-3.47) | 0.836 |
| Insertive only | **3.33(1.27-8.69)** | **0.014** | 1.98(0.66-5.91) | 0.221 | 2.01(0.59-6.75) | 0.263 |
| **Biological and health characteristics** |  |  |  |  |  |  |
| **Mental Health status** |  |  |  |  |  |  |
| No depressive symptoms | Ref |  | Ref |  | Ref |  |
| Depressed | **2.64(1.51-4.61)** | **0.001** | **2.09(1.08-4.06)** | **0.028** | 1.25(0.47-3.35) | 0.658 |
| **Self-reported STI history in the 12 months prior to the study** |  |  |  |  |  |  |
| No | Ref |  | Ref |  | Ref |  |
| Yes | **1.68(1.12-2.52)** | **0.012** | **1.63(1.01-2.63)** | **0.045** | 2.18(0.94-5.08) | 0.07 |
| **HIV infection** |  |  |  |  |  |  |
| Negative | Ref |  |  |  |  |  |
| Positive | 0.93(0.52-1.65) | 0.802 |  |  |  |  |
| **STI infection** |  |  |  |  |  |  |
| Negative | Ref |  |  |  |  |  |
| Positive | 1.26(0.83-1.92) | 0.272 |  |  |  |  |
| **Urethral STI** |  |  |  |  |  |  |
| Negative | Ref |  |  |  |  |  |
| Positive | 0.78(0.39-1.53) | 0.467 |  |  |  |  |
| **Rectal STI** |  |  |  |  |  |  |
| Negative | Ref |  | Ref |  | Ref |  |
| Positive | **2.26(1.26-4.06)** | **0.006** | **2.65(1.35-5.23)** | **0.005** | **4.19(2.02-8.72)** | **0.0001** |

### Testing outcomes of self-collected rectal swabs

Overall, 10% (75/738) of rectal swab samples gave an indeterminate result on the first attempt and the test had to be repeated on a fresh sample. These included 65 invalid results and 10 errors. Overall, 21%(16/65) of participants who had an indeterminate result the first time had to repeat the specimen collection (11 participants repeated the test twice and 5 participants repeated the test three times). Upon retesting, 85% (64/75) of participants with initial indeterminate results tested negative, 12% (9/75) tested positive for CT or NG, only 2 samples remained indeterminate.

Young MSM/TGW were more likely to have an indeterminate result compared to older MSM/TGW (χ2 P: 0.011). Proportions of MSM/TGW with indeterminate results were 13% (45/338) among 18–24-year-olds, 9% (26/296) among 25-34 and 4% (4/104) among participants aged ≥ 35. Participants with STI symptoms in the previous 12 months were also less likely to have an indeterminate result compared to those who did not have symptoms (5% vs 11%, χ2 P: 0.033). However, there was no difference based on any behavioral and biological factors.

## Discussion

MSM and TGW in Kigali, Rwanda were able to collect patient-collected rectal swabs for STI testing with minimal guidance, and most reported that collection was easy, comfortable, and highly acceptable. Given the high prevalence of asymptomatic rectal infections among MSM and TGW in Rwanda, introduction of self-collection of rectal samples could be an additional tool to support routine STI screening in Rwanda.

Self-collected biological specimen for STI testing, including rectal swabs, have been shown to have similar diagnostic accuracy as provider-collected samples in many studies ^9,15,30,31^. Moreover, self-collection offers a less intrusive alternative for extragenital screening among MSM and TGW, many of whom may be reluctant to undergo provider-administered testing. As such, self-collection of biological specimens including rectal swabs has successfully been integrated into clinical settings ^13,14,26^. Self-collection extends opportunities for STI testing outside of clinical settings, which further justifies their integration in STI programming in Rwanda. For instance, self-collection of rectal swabs could support community-based STI testing among MSM/TGW who have limited access to culturally competent clinic-based services. Community provision of HIV and STI services for MSM and TGW are recommended by the World Health Organization ^32^. Furthermore, self-collection of rectal swabs can support home-based collection. Several studies have shown home-based sample collection to be acceptable, convenient, and timesaving ^15,16^. Home-based collection could prove beneficial as it can improve partner notification, testing, and treatment ^33^. Finally, although evidence is still limited, self-collection of biological swabs seems to be a cost-effective option compared to sample collection by healthcare providers. For example, a study in the UK found that self-collected swabs were less expensive than clinician-collected swabs in clinic or community settings where asymptomatic testing is being performed ^34^. Similarly, in a study in Australia of a home-based STI retesting strategy using self-collection kits, the overall cost for home retesting was lower than clinic retesting ^35^. In Africa, several home-based testing interventions have been implemented and successfully improved testing and linkage to care for various infections including HIV, STI and tuberculosis ^36-38^. However, in Africa, home testing is usually conducted by community health workers or healthcare providers and not beneficiaries themselves. All these advantages make self-collection of rectal swabs an option worth exploring in Rwanda and other countries where STI testing is still challenging.

Results from this study highlight that one in seven participants found the self-collection of the rectal swab to be difficult. Notably, participants with a history of STI and with prevalent rectal STI were more likely to find rectal swabbing difficult. This is particularly challenging programmatically, because these individuals would benefit the most from any screening/testing programs to avoid further complications or risks associated with STI. We also found that discomfort with the test was significantly associated with difficulty in self-collecting the rectal specimen. Despite these findings, almost all participants reported that they would consider repeat self-collection, suggesting that discomfort might not be a major barrier to future self-collection. However, these findings demonstrate the need to improve the overall experience of self-collection of rectal swabs in Rwanda for some men, especially those with STI. For instance, the provision of instructions on sample collection could further be improved especially for people who are more likely to find the procedure difficult or uncomfortable. In our study, participants were verbally instructed on methods of self-collection by trained nurses, but visual instructional strategies, including printed pictorial instructions, or the use of culturally relevant videos have been employed in other studies^10,16,39^. Development of such tools that are adapted to Rwandan context could improve rectal swab self-collection experience. Further research including qualitative and empirical research could help explain the reasons behind experiencing the difficulty/discomfort with the procedure and propose solutions. Finally, educational efforts aiming to increase awareness and reduce the stigma associated with the test among providers and/or MSM/TGW individuals are needed.

These data also raise important considerations for any program trying to integrate self-collection of rectal swabs among MSM and TGW within and outside clinical settings in Rwanda. For instance, 10% of samples yielded indeterminate results, and some participants had to repeat the test several times. This carries significant cost and clinical implications. Clinical consequences include delayed diagnosis and treatment, which may result in preventable STI complications or downstream transmission to sexual partners. Repeat testing also carries cost implications. However, given the lower cost of self-collection compared to clinician-collected testing, the additional cost of repeat testing might still be less than paying clinicians to collect the tests. There are also additional logistical challenges that may arise from STI testing in the community or home settings including appropriate sample storage and transport systems for samples collected outside of clinical settings. These logistical considerations influence turnaround time and test performance. Additionally, programs will need to design notification systems to ensure timely treatment of individuals diagnosed with STI and their infected partners as well as retesting after indeterminate samples. For this, point-of-care tests that fulfill the ASSURED criteria - affordable, sensitive, specific, user-friendly, robust/rapid, equipment-free, and delivered to end-users - criteria could support these programs.^40^ Finally, the costs implications of such programs need to be balanced with their potential benefits, especially for countries with limited financial resources. All these challenges demonstrate the need for carefully designed implementation science studies to ensure the successful implementation of rectal swab self-collection in clinics, communities, and home settings in Rwanda and other LMICs. These studies should include a comprehensive costing of integrating self-collection of rectal swabs, urine and oropharyngeal specimen in clinics, community- and home settings; to allow for detailed cost-effectiveness analyses comparing this intervention to the standard of care.

This study has some limitations. Willingness to have biological testing, including a rectal swab, was an eligibility criterion of the study. However, refusal of biological samples was only expressed by < 20% of the 157 individuals who were ineligible. Because of this selection bias, the high willingness to use the self-collected test in the future found in this study might be an overestimate of the actual acceptability in the larger population of MSM/TGW in Kigali. We are also unable to discuss the reasons why the proportion of indeterminate results in our study is higher than what has been reported in other studies ^41,42^. Indeterminate results of rectal swabs may occur for various reasons including the presence of fecal material, mucus, and blood in the sample, among others ^43^. However, we did not collect this information or other information about rectal hygiene practices. Thus, any future studies leveraging self-collection of rectal swabs for STI testing in Rwanda also should try to systematically collect data on potential causes of indeterminate results when feasible. Future studies also should investigate why young MSM/TGW seem to have a higher occurrence of indeterminate results. Another limitation of our study is the lack of data comparing the diagnostic accuracy of self-collected samples versus clinician-collected specimens specific to Rwanda. Finally, the results are subject to social desirability bias, because the interviewers who provided the instructions and sample collection kits were the same people collecting data on participants’ experiences with these tests. Despite these limitations, our data provide important information to guide future rectal STI testing strategies in Rwanda and other LMICs.

## Conclusion

Self-collection of rectal specimens for STI testing was highly acceptable and yielded valid biological samples for the vast majority participants in this sample of MSM/TGW from Kigali, Rwanda. Integration of self-collection of biological specimens could be introduced as a component of STI programming for MSM/TGW in Rwanda. For optimal results, carefully designed implementation science studies will be needed to inform the sample collection procedures, instructions provided to clients, specimen transportation, provision of results to patients, and treatment and follow up both in clinical and community settings.

## Data Availability

All data produced in the present study are available upon reasonable request to the authors

## Authors’ contributions

SDB, SA, EK, PS, AK conceived and designed the study. JOTR, BL, CEL oversaw implementation and data collection. JOTR analyzed the data and wrote the first draft of the paper. All authors reviewed, edited, and approved the manuscript.

## Acknowledgements

The authors thank all members of MSM and transgender individuals operating in Kigali city who helped during the study design and implementation. We would also like to thank all the study participants for their time, commitment, and contribution to this study

## List of abbreviations

ART: Antiretroviral treatment
CCU: Consistent Condom Use
CT: Chlamydia Trachomatis
ELISA: Enzyme-linked Immunosorbent Assay
Frw: Francs Rwandais
HDI: Health Development Initiative
HIV: Human Immunodeficiency virus
IQR: Interquartile range
MSM: men who have sex with men
MSMW: men who have sex with men and women
NG: Neisseria Gonorrhea
NRL: National reference Laboratory of Rwanda
PrEP: Pre-exposure Prophylaxis
PSF: Projet San Francisco
RDS: Respondent Driven Sampling
RPR: Rapid Plasma Reagin
SSA: Sub-Saharan Africa
STI: Sexually Transmitted Infection
TGW: Transgender women
TPHA: Treponema Pallidum Hemagglutination Assay
UNAIDS: Joint United Nations Programme on HIV/AIDS

## References

1. James SL, Abate D, Abate KH, et al. Global, regional, and national incidence, prevalence, and years lived with disability for 354 diseases and injuries for 195 countries and territories, 1990–2017: a systematic analysis for the Global Burden of Disease Study 2017. The Lancet. 2018;392(10159):1789–1858.

2. Katz DA, Dombrowski JC, Bell TR, Kerani RP, Golden MR. HIV Incidence Among Men Who Have Sex With Men After Diagnosis With Sexually Transmitted Infections. Sex Transm Dis. 2016;43(4):249–254.

3. Unemo M, Bradshaw CS, Hocking JS, et al. Sexually transmitted infections: challenges ahead. The Lancet Infectious Diseases. 2017;17(8):e235–e279.

4. Baral S, Sifakis F, Cleghorn F, Beyrer C. Elevated risk for HIV infection among men who have sex with men in low- and middle-income countries 2000-2006: a systematic review. PLoS Med. 2007;4(12):e339.

5. Mayer KH, Allan-Blitz LT. Similar, but different: drivers of the disproportionate HIV and sexually transmitted infection burden of key populations. J Int AIDS Soc. 2019;22 Suppl 6:e25344.

6. Baral SD, Grosso A, Holland C, Papworth E. The epidemiology of HIV among men who have sex with men in countries with generalized HIV epidemics. Current Opinion in HIV and AIDS. 2014;9(2):156–167.

7. Twahirwa Rwema JO, Herbst S, Hamill MM, et al. Cross-sectional assessment of determinants of STIs among men who have sex with men and transgender women in Kigali, Rwanda. Sexually Transmitted Infections. 2021:sextrans-2020-054753.

8. Patton ME, Kidd S, Llata E, et al. Extragenital gonorrhea and chlamydia testing and infection among men who have sex with men--STD Surveillance Network, United States, 2010-2012. Clin Infect Dis. 2014;58(11):1564–1570.

9. van der Helm JJ, Hoebe CJ, van Rooijen MS, et al. High performance and acceptability of self-collected rectal swabs for diagnosis of Chlamydia trachomatis and Neisseria gonorrhoeae in men who have sex with men and women. Sex Transm Dis. 2009;36(8):493–497.

10. Sexton ME, Baker JJ, Nakagawa K, et al. How reliable is self-testing for gonorrhea and chlamydia among men who have sex with men? Journal of Family Practice. 2013;62(2):70–78.

11. Moncada J, Schachter J, Liska S, Shayevich C, Klausner JD. Evaluation of self-collected glans and rectal swabs from men who have sex with men for detection of Chlamydia trachomatis and Neisseria gonorrhoeae by use of nucleic acid amplification tests. J Clin Microbiol. 2009;47(6):1657–1662.

12. Alexander S, Ison C, Parry J, et al. Self-taken pharyngeal and rectal swabs are appropriate for the detection of Chlamydia trachomatis and Neisseria gonorrhoeae in asymptomatic men who have sex with men. Sex Transm Infect. 2008;84(6):488–492.

13. Barbee LA, Tat S, Dhanireddy S, Marrazzo JM. Implementation and Operational Research: Effectiveness and Patient Acceptability of a Sexually Transmitted Infection Self-Testing Program in an HIV Care Setting. J Acquir Immune Defic Syndr. 2016;72(2):e26–31.

14. Soni S, White JA. Self-screening for Neisseria gonorrhoeae and Chlamydia trachomatis in the human immunodeficiency virus clinic--high yields and high acceptability. Sex Transm Dis. 2011;38(12):1107–1109.

15. Wayal S, Llewellyn C, Smith H, et al. Self-sampling for oropharyngeal and rectal specimens to screen for sexually transmitted infections: acceptability among men who have sex with men. Sex Transm Infect. 2009;85(1):60–64.

16. John SA, Cain D, Bradford-Rogers J, Rendina HJ, Grov C. Gay and bisexual men’s experiences using self-testing kits for HIV and rectal and urethral bacterial sexually transmitted infections: Lessons learned from a study with home-based testing. Int J Sex Health. 2019;31(3):308–318.

17. Twahirwa Rwema JO, Lyons CE, Herbst S, et al. HIV infection and engagement in HIV care cascade among men who have sex with men and transgender women in Kigali, Rwanda: a cross-sectional study. Journal of the International AIDS Society. 2020;23(S6).

18. Adedimeji A, Sinayobye JD, Asiimwe-Kateera B, et al. Social contexts as mediator of risk behaviors in Rwandan men who have sex with men (MSM): Implications for HIV and STI transmission. PLoS One. 2019;14(1):e0211099.

19. Organization WH. Progress report on Implementation of the Global Strategy for Prevention and Control of Sexually Transmitted Infections: 2006–2015. plGeneva: World Health Organization;2015.

20. Grijsen ML, Graham SM, Mwangome M, et al. Screening for genital and anorectal sexually transmitted infections in HIV prevention trials in Africa. Sex Transm Infect. 2008;84(5):364–370.

21. Zemouri C, Wi TE, Kiarie J, et al. The Performance of the Vaginal Discharge Syndromic Management in Treating Vaginal and Cervical Infection: A Systematic Review and Meta-Analysis. PLoS One. 2016;11(10):e0163365.

22. Jones J, Sanchez TH, Dominguez K, et al. Sexually transmitted infection screening, prevalence and incidence among South African men and transgender women who have sex with men enrolled in a combination HIV prevention cohort study: the Sibanye Methods for Prevention Packages Programme (MP3) project. J Int AIDS Soc. 2020;23 Suppl 6:e25594.

23. Scheim AI, Travers R. Barriers and facilitators to HIV and sexually transmitted infections testing for gay, bisexual, and other transgender men who have sex with men. AIDS Care. 2017;29(8):990–995.

24. Kim H-Y, Grosso A, Ky-Zerbo O, et al. Stigma as a barrier to health care utilization among female sex workers and men who have sex with men in Burkina Faso. Annals of Epidemiology. 2018;28(1):13–19.

25. Wiginton JM, Murray SM, Poku O, et al. Disclosure of same-sex practices and experiences of healthcare stigma among cisgender men who have sex with men in five sub-Saharan African countries. BMC Public Health. 2021;21(1):2206.

26. Ogale Y, Yeh PT, Kennedy CE, Toskin I, Narasimhan M. Self-collection of samples as an additional approach to deliver testing services for sexually transmitted infections: a systematic review and meta-analysis. BMJ Glob Health. 2019;4(2):e001349.

27. Tordoff DM, Morgan J, Dombrowski JC, Golden MR, Barbee LA. Increased Ascertainment of Transgender and Non-binary Patients Using a 2-Step Versus 1-Step Gender Identity Intake Question in an STD Clinic Setting. Sex Transm Dis. 2019;46(4):254–259.

28. Kroenke K, Spitzer RL, Williams JBW. The PHQ-9. Journal of General Internal Medicine. 2001;16(9):606–613.

29. Okonkwo N, Rwema JOT, Lyons C, et al. The Relationship Between Sexual Behavior Stigma and Depression Among Men Who have Sex with Men and Transgender Women in Kigali, Rwanda: a Cross-sectional Study. International Journal of Mental Health and Addiction. 2021.

30. Lunny C, Taylor D, Hoang L, et al. Self-Collected versus Clinician-Collected Sampling for Chlamydia and Gonorrhea Screening: A Systemic Review and Meta-Analysis. PLoS One. 2015;10(7):e0132776.

31. Freeman AH, Bernstein KT, Kohn RP, Philip S, Rauch LM, Klausner JD. Evaluation of self-collected versus clinician-collected swabs for the detection of Chlamydia trachomatis and Neisseria gonorrhoeae pharyngeal infection among men who have sex with men. Sex Transm Dis. 2011;38(11):1036–1039.

32. Who. Consolidated guidelines on HIV prevention, diagnosis, treatment and care for key populations. 2016.

33. Andersen B, Østergaard L, Møller JK, Olesen F. Home sampling versus conventional contact tracing for detecting <em>Chlamydia trachomatis</em> infection in male partners of infected women: randomised study. BMJ. 1998;316(7128):350–351.

34. Wilson JD, Wallace HE, Loftus-Keeling M, et al. Swab-yourself trial with economic monitoring and testing for infections collectively (SYSTEMATIC): Part 2. A diagnostic accuracy, and cost-effectiveness, study comparing rectal, pharyngeal and urogenital samples analysed individually, versus as a pooled specimen, for the diagnosis of gonorrhoea and chlamydia. Clin Infect Dis. 2020.

35. Smith KS, Kaldor JM, Hocking JS, et al. The acceptability and cost of a home-based chlamydia retesting strategy: findings from the REACT randomised controlled trial. BMC Public Health. 2016;16:83.

36. Pathmanathan I, Nelson R, de Louvado A, et al. High Coverage of Antiretroviral Treatment With Annual Home-Based HIV Testing, Follow-up Linkage Services, and Implementation of Test and Start: Findings From the Chokwe Health Demographic Surveillance System, Mozambique, 2014-2019. J Acquir Immune Defic Syndr. 2021;86(4):e97–e105.

37. Uwimana J, Zarowsky C, Hausler H, Jackson D. Training community care workers to provide comprehensive TB/HIV/PMTCT integrated care in KwaZulu-Natal: lessons learnt. Trop Med Int Health. 2012;17(4):488–496.

38. Koduah Owusu K, Adu-Gyamfi R, Ahmed Z. Strategies To Improve Linkage To HIV Care In Urban Areas Of Sub-Saharan Africa: A Systematic Review. HIV AIDS (Auckl). 2019;11:321–332.

39. Lampinen TM, Latulippe L, van Niekerk D, et al. Illustrated instructions for self-collection of anorectal swab specimens and their adequacy for cytological examination. Sex Transm Dis. 2006;33(6):386–388.

40. Wi TE, Ndowa FJ, Ferreyra C, et al. Diagnosing sexually transmitted infections in resource-constrained settings: challenges and ways forward. J Int AIDS Soc. 2019;22 Suppl 6:e25343.

41. Parcell BJ, Ratnayake L, Kaminski G, Olver WJ, Yirrell DL. Value of repeat testing using Cepheid GeneXpert CT/NG for indeterminate PCR results when diagnosing Chlamydia trachomatis and Neisseria gonorrhoeae. Int J STD AIDS. 2015;26(1):65–67.

42. Gaydos CA, Van Der Pol B, Jett-Goheen M, et al. Performance of the Cepheid CT/NG Xpert Rapid PCR Test for Detection of Chlamydia trachomatis and Neisseria gonorrhoeae. J Clin Microbiol. 2013;51(6):1666–1672.

43. Schrader C, Schielke A, Ellerbroek L, Johne R. PCR inhibitors - occurrence, properties and removal. J Appl Microbiol. 2012;113(5):1014–1026.

